# A random-walk-based epidemiological model

**DOI:** 10.1101/2020.11.04.20226308

**Authors:** Andrew Chu, Greg Huber, Aaron McGeever, Boris Veytsman, David Yllanes

## Abstract

Random walkers on a two-dimensional square lattice are used to explore the spatio-temporal growth of an epidemic. We have found that a simple random-walk system generates non-trivial dynamics compared with traditional well-mixed models. Phase diagrams characterizing the long-term behaviors of the epidemics are calculated numerically. The functional dependence of the basic reproductive number *R*_0_ on the model’s defining parameters reveals the role of spatial fluctuations and leads to a novel expression for *R*_0_. Special attention is given to simulations of inter-regional transmission of the contagion. The scaling of the epidemic with respect to space and time scales is studied in detail in the critical region, which is shown to be compatible with the directed-percolation universality class.

## Introduction

Classic epidemiological models^1, 2^ often assume panmictic populations: every infected person has an equal chance to affect any other person in the population. While these models have been successful in describing an ideal dynamics of epidemic spread, they do not account for inhomogeneity: an infected person has higher probability to transmit the disease to a member of their household^3, 4^ or to a person in their locale. The overall reduction of long-distance travel due to the COVID-19 pandemic makes the latter cause of inhomogeneity especially relevant: many infected persons (but not all) spread the infection only in their immediate vicinity. Likewise, the local neighborhoods of both susceptible and infected persons are dynamic – they vary in space and time depending on the local state of the epidemic. These spatio-temporal inhomogeneities are studied in this paper.

Research into models of spatial epidemiology have produced a number of approaches to the limitations of well-mixed models.^5–9^ These works range from exploring known social networks^7^ to looking at possible policy responses to the current pandemic.^5^ The current paper focuses on a simple random-walk model that lends itself to analytical analysis, in order to explore epidemic properties that arise in the presence of simple spacial dynamics.

There are two approaches to geographical inhomogeneity: detailed analyses based on contact-tracing and mobility data,^10, 11^ or analyses based on stylized models.^4, 12, 13^ The first approach, while potentially highly accurate, requires a large number of parameters and is not robust with respect to unforeseen changes in mobility patterns. The second approach, in contrast, requires a small number of well-defined parameters and provides a sound intuition about their effects on the outcome. It is especially useful when planning interventions and policy changes, or investigating various “what if” scenarios. It is the latter approach that we have taken here.

In the present paper, we treat a very simple model: a random walker (modeled on an infected person) takes *τ* steps (a positive integer) on a 2D square lattice. After each step, the visited site can become infected with probability *p*. In this case, at the site of the new infection, it branches off another random walker, also with lifetime *τ*, that can also further spread the contagion to further sites, and so on. We study the resulting epidemic in space and time.

This model depends on two parameters: *τ*, representing the length of time of the infectious period, and *p*, representing the infectiousness. It also depends on the “walking pattern”, which may include steps to the neighboring sites or jumps to more distant locations.

Panmictic models predict that the spread of epidemics depends only on the basic reproduction number *R*_0_, the number of infected persons produced by one walker in a fully susceptible population, which, in turn, can be estimated in the panmictic approximation as

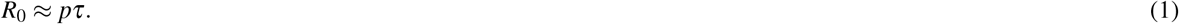

We will refer to this estimate as the *naïve approximation*.

However, the random-walk model exhibits a surprisingly rich behavior, well beyond the “naïve approximation” above. We will show that the spread of the epidemics depends on the interplay between *p* and *τ*. Moreover, the detailed geometry of the epidemics depends on the stochasticity of individual walks and the nature of random walks on the plane. In subsequent sections, we describe our model in full, study its behavior using numerical simulations and theory, and draw conclusions for real-world epidemics.

## Model

We model the geographic distribution of the population by an infinite 2D square lattice. At any given time, each site can be in one of three states. Although the model proposed here is fundamentally different than an SIR model, the same nomenclature is adopted; namely, each site in the lattice is either *susceptible, infected*, or *removed*. (Figure 1). All sites are initialized as susceptible, with the exception of the origin which contains a single infectious agent. An infected site generates a brand new random walker, which starts to move on the lattice. If the walker lands on a susceptible site (sites are represented as square cells in the figure), it infects it with probability *p*. The infected site starts a new walker and itself becomes removed (no longer susceptible to infection). After *τ* steps, the walker recovers and no longer infects other sites. The label “removed” is used in the same manner as in well-mixed SIR models; these sites are not physically removed from the lattice but instead removed from the population of susceptible sites and can no longer be infected.

**Figure 1.**
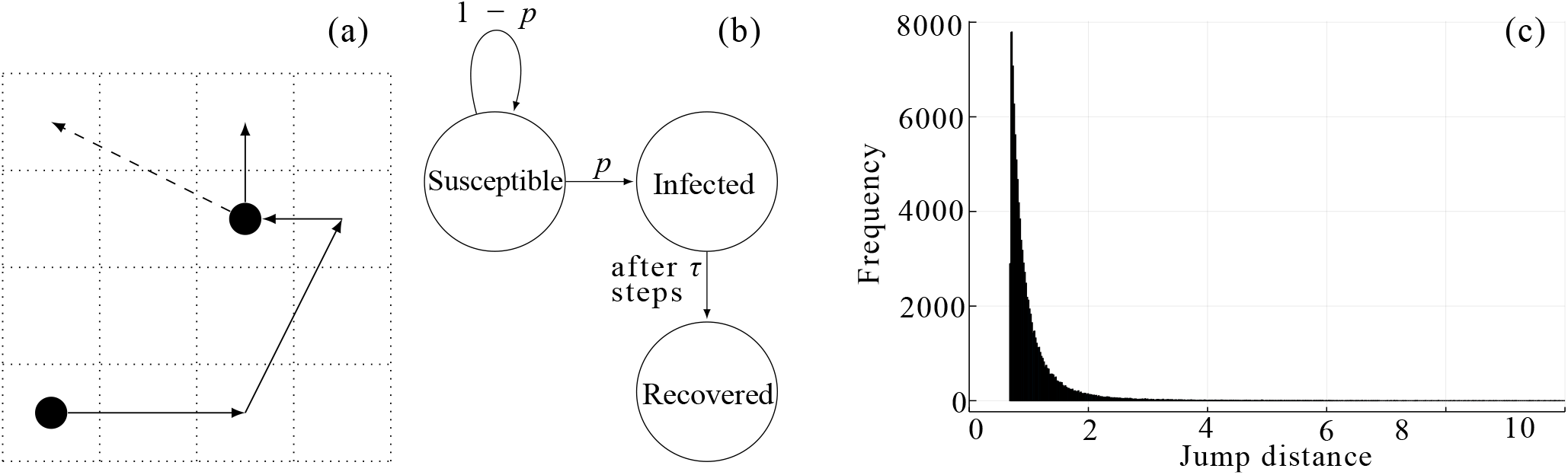
The model. (a) A walker (solid line) at some step infects a susceptible site, which generates another walker (dashed line). (b) The walker state diagram for each step, where *p* and 1 − *p* transitions are triggered by infectious-susceptible interactions. (c) Histogram of jump distances of infective agents for 100000 realizations.

Thus, there are two fundamental parameters that control the outbreak: *p*, the probability of infection; and *τ*, the number of steps for which an infective agent (walker) is active before recovering.

Another feature of the model is the way the infectious agent chooses its next move. The model is quasilocal: most of the time the walker moves to one of the eight adjacent sites, but sometimes it makes a longer jump. Namely, if the walker is at **r**_0_ = (*x*_0_, *y*_0_), its next location, **r**, is given by

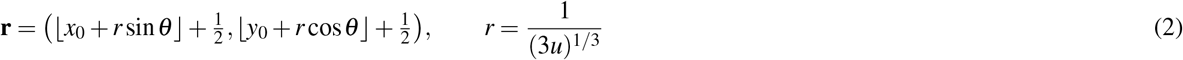

with *u* and *θ* drawn from uniform distributions:

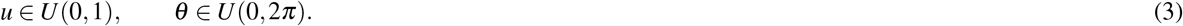

Here ⌊*X*⌋ is the integer part of *X* (the largest integer number not exceeding *X*), and *U* (*a,b*) is the uniform distribution in the interval [*a,b*] —the nodes of the lattice have integer coordinates, but our walkers move on the dual lattice, consisting of the centers of the elementary cells—. This function produces a one-step walk to one of the eight adjacent sites approximately 66.8% of the time; 95.7% of jumps are within a distance of 2 sites. The random-variate generator for the jump distances *r* in eq. (2) is derived from a probability density function of the form *ϕ*(*r*) = *A/r*^4^, where the constant *A* is of order the lattice constant cubed *a*^3^ (in this paper, *a* = 1 is chosen throughout). Our choice of this kernel seeks a balance, allowing for discrete jumps and generating more interesting dynamics than simple diffusion, while avoiding a regime dominated by long-range jumps (because we focus on the short-range evolution of a local outbreak, not on the dynamics caused by, e.g., long-range travel). In fact, the exponent of —4 corresponds to interactions of the form ∼1*/r*^*d*+*σ*^ when *d* = 2, *σ* = 2, a case of special interest in a number of early studies,^14, 15^ because it sits at the margin between long-range and short-range interactions. However, in the context of epidemic models, as noted by Bunde *et al*. ^16^, Grassberger ^12^, and Hallatschek and Fisher ^13^, it presents no special difficulties.

One interpretation of this model is that an infectious agent is a representative of a tight-knit population, household or community.^3, 4^ The susceptible communities are dense and fill the entire lattice. Each time an infection occurs, a distribution of possible further propagation in neighboring communities is generated, which we realize by considering a random walk by a representative infectious agent from the community. The originating community is then removed from the susceptible pool, although visitations are still allowed, which can be viewed as coming from several causes. The community could undergo lockdown/quarantine measures after the initial infections and no further spread occurs. Or, the epidemic burns through the community leaving only recovered members, so the community is no longer susceptible.

## Results

### Initial stages of the epidemics

In the initial stage of the outbreak, the infected person (the index case) is completely surrounded by the susceptible population. The key parameter describing this state is *R*_0_: the number of people infected by the index case. The value of *R*_0_ depends on *p* and *τ* in some form, but the naïve approximation, *R*_0_ ≈ *pτ*, must fail, as can be seen whenever the path of even a single walker self intersects; *i*.*e*., the approximation does not account for the recurrence of a two-dimensional random walker.

We ran 100000 realizations of the simulation, collecting how many agents were infected by the index case. Table 1 shows the *R*_0_ calculated for *p* and *τ* tabulated on log-log axes, an idea that will be explored further in the discussion of the phase diagram. Notice that *R*_0_ is slightly less than 1 for *p* = *τ* = 1 because there is a very small probability that the walker will not leave its starting cell. The breakdown of the naïve approximation is evident in the lack of constancy of *R*_0_ along the diagonals of the table, which is particularly apparent when *pτ* is large.

**Table 1.** *R*_0_ values calculated over 2500000 realizations. The numbers in parenthesis are the standard error in the last digits. *R*_0_ values which lie in the regime of indefinite spreading are bolded.

| $\tau \backslash p$ | 1 | 1/2 | 1/4 | 1/8 | 1/16 | 1/32 | 1/64 |
| --- | --- | --- | --- | --- | --- | --- | --- |
| 1 | 0.99927(2) | 0.4997(3) | 0.2498(3) | 0.1252(2) | 0.0627(2) | 0.03130(11) | 0.01577(8) |
| 2 | **1.8614(2)** | 0.9307(4) | 0.4658(4) | 0.2339(3) | 0.1171(2) | 0.0583(2) | 0.02896(11) |
| 3 | **2.67771(0)** | 1.37355(0) | 0.69514(0) | 0.34982(0) | 0.17546(0) | 0.08794(0) | 0.044(0) |
| 4 | **3.4590(4)** | **1.8042(6)** | 0.9211(5) | 0.4658(4) | 0.2339(3) | 0.1171(2) | 0.0586(2) |
| 5 | **4.21565(0)** | **2.23352(0)** | 1.14976(0) | 0.58241(0) | 0.29344(0) | 0.14692(0) | 0.0736(0) |
| 8 | **6.3862(7)** | **3.4885(8)** | **1.8251(7)** | 0.9334(6) | 0.4723(4) | 0.2375(3) | 0.1189(2) |
| 10 | **7.77848(0)** | **4.30866(0)** | **2.27345(0)** | 1.16819(0) | 0.59184(0) | 0.29747(0) | 0.14961(0) |
| 16 | **11.7767(13)** | **6.7137(11)** | **3.6020(10)** | **1.8686(8)** | 0.9512(6) | 0.4802(4) | 0.2414(3) |
| 32 | **21.725(2)** | **12.8762(16)** | **7.0810(14)** | **3.7223(11)** | **1.910(8)** | 0.9677(6) | 0.4870(4) |
| 64 | **40.214(3)** | **24.643(2)** | **13.8614(19)** | **7.3885(15)** | **3.8184(12)** | **1.9432(9)** | 0.9793(6) |
| 128 | **74.711(6)** | **47.143(3)** | **27.092(2)** | **14.629(2)** | **7.6183(16)** | **3.8883(12)** | **1.9663(9)** |

The theoretical result (see Methods) is

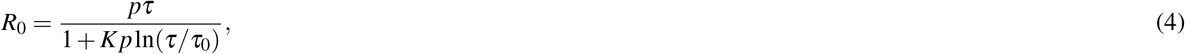

where *K* and *τ*_0_ are constants that depend on how the walker’s next jump is selected. For the kernel in equation (2) we have

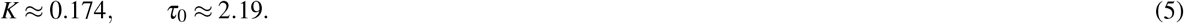

As shown in Figure 2, this formula is in excellent agreement with the numerical experiments without any adjustable parameters.

**Figure 2.**
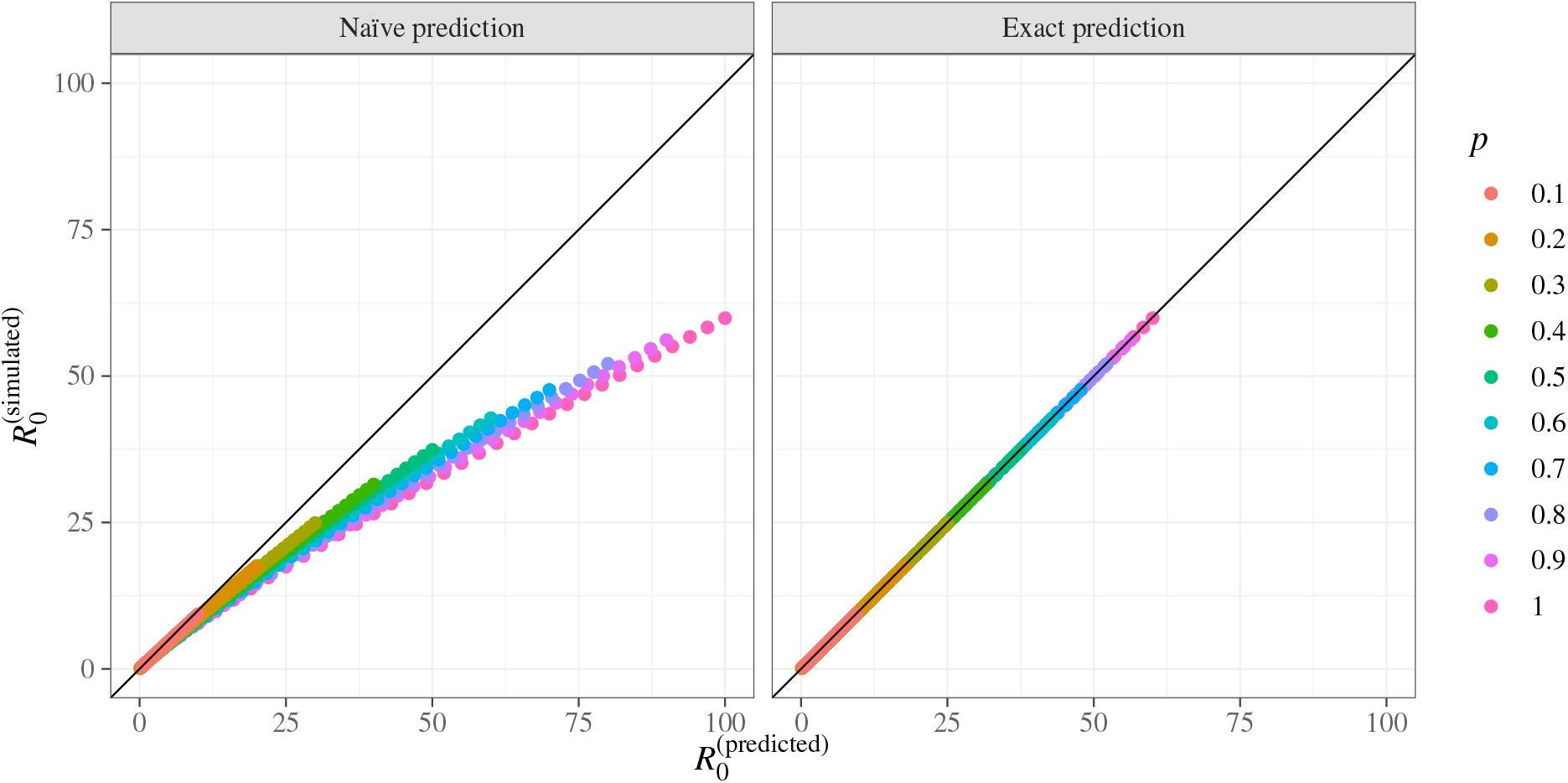
The prediction for *R*_0_ and simulation data. Left panel: actual 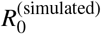 against the naïve prediction of Eq, ((1)), 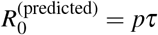. Right panel: 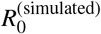 against the prediction of eq. (4). The straight line is 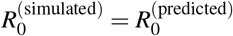. Each dot corresponds to a combination of *p* and *τ* (the values of *p* are color coded, *τ* is between 1 and 100).

An interesting question is the interplay between the length of the walk and the jump length: what is more important for the epidemic’s dynamics, the length of the infectious period or the span of contacts of the infected individuals? As shown in the Methods section, the coefficients in equation (4) scale as *K* ∝ *c*^−1^, *τ*_0_ ∝ *c*^−1^, where *c* is the coefficient of the Fourier expansion of the structure factor. The dimension of *c* is squared length, so we can write down *c* ∝ *ℓ*^2^, where *ℓ* is the average jump length, or, in other words, the spread of contacts for an average individual. Equation (4) shows that for small infectivity *p* the result does not depend on *ℓ*. However, for large *p*, when *Kp*ln(*τ/τ*_0_) ≫ 1, the number *R*_0_ depends on *τℓ*^2^, *i*.*e*., the number of persons infected by a single individual scales as the square of the jump length (and linearly with the number of jumps). This conclusion might inform the policies for outbreak suppression and the details of stay-at-home orders.

### Outbreak progression and phase diagrams

The progression of the outbreak beyond the initial stage is illustrated by Figure 3. Depending on the parameters, the outbreak may die out or spread indefinitely, and it is a natural starting point to describe this dependence by a phase diagram.

**Figure 3.**
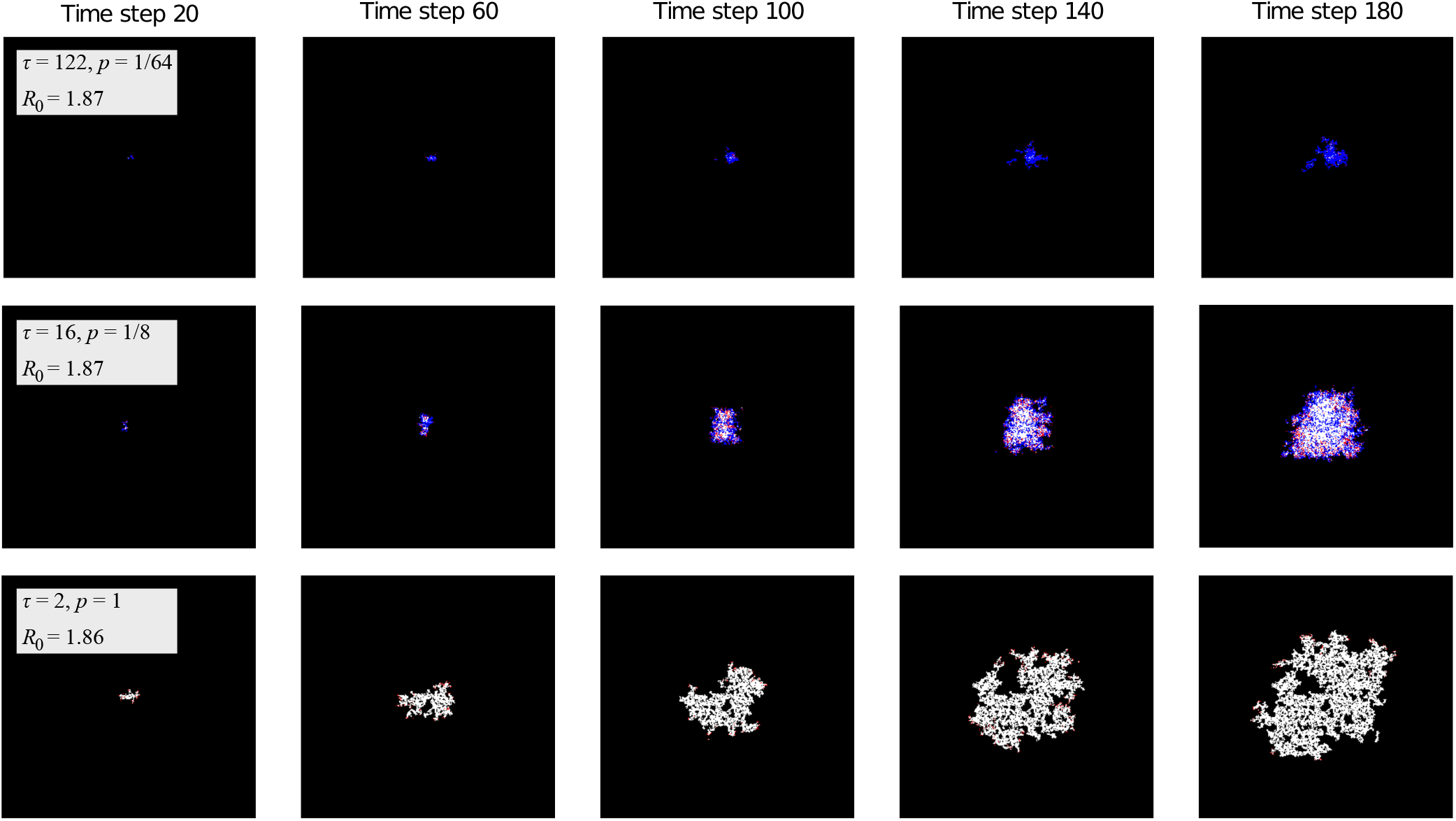
Progression of the outbreak. Red sites are infective agents, white are removed and black are susceptible. Blue sites are not technically part of the SIR categories but represent susceptible sites which have been previously visited (but not infected). This fourth color brings attention to the pockets of susceptibles left in the wake of the infection. Note that the model exhibits very different behavior for combinations of *τ* and *p* with almost identical *R*_0_, demonstrating that *R*_0_ is not always an accurate characterization of the overall behavior of an outbreak. For example, the outbreak in the bottom row grew much more rapidly in the earlier time steps, but by the final time step, the outbreak in the middle row had a much larger population of active infective agents. Nonetheless, the cumulative number of infections is much higher in the former.The outbreak shown in the top row is clearly of much smaller magnitude despite having the same *R*_0_. The boxes are 300 × 300 sites.

Phase diagrams are used in physics and related fields as a visualization of how pressure and temperature (or other thermodynamic variables) affect the bulk thermodynamic state of a substance. Here, each point in the phase diagram will describe the expected behavior of an epidemic with the given *p* and *τ* inputs (*i*.*e*., how likely it is to die out after a certain amount of time has passed).

For each (*p, τ*) pair we launched many simulations and used to define the *phase* of an infection by the following metric [see Supplemental Material (SM) for details]:

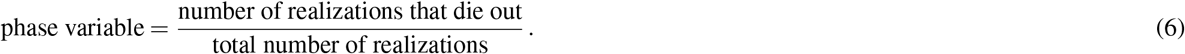

The plot of this phase variable is shown in Figure 4. The determination of the phase boundary allows us to predict the behavior of the system in the regions above or below the boundary, even in the absence of extensive simulations. For each horizontal cross-section of the phase diagram (corresponding to a 1D phase diagram for a fixed value of *τ*), we have fitted the phase variable as a function of *p* to a smooth interpolating function in order to find the location of the phase boundary, defined as the point where the phase variable is 0.5. The resulting phase boundary is well approximated by a hyperbola in the *p, τ* plane. In particular, a least-squares fit gives the curve 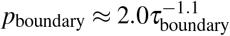.

**Figure 4.**
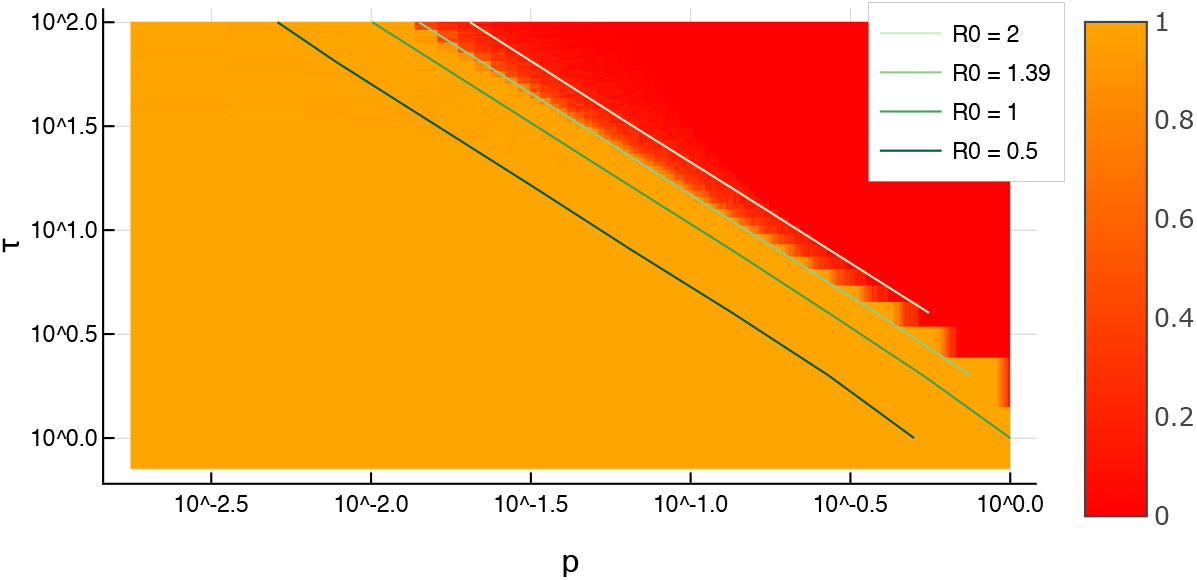
Phase diagram in log-log scale, including iso-*R*_0_ lines. The heatmap scale shows the phase variable of Eq. (6), namely, the fraction of realizations where the outbreak dies out. While a continuous red-orange color scale is used to plot the phase variable, the phase diagram appears as essentially a binary plot of two colors. This behavior reflects the sharp transition.

It is customary in epidemiology^2^ to predict the progression of epidemics based on the value of *R*_0_. Accordingly, on Figure 4 “iso-*R*_0_” lines were overlaid on the phase diagram: each point on an iso-*R*_0_ line has *p* and *τ* values which produce outbreaks with a common *R*_0_. As expected, the phase boundary indeed coincides with an iso-*R*_0_ line, having *R*_0_ ≈ 1.39 (not *R*_0_ = 1, because of the role of statistical fluctuations). As shown in the Methods section, this finding corresponds to a robust theoretical prediction. Hence, while two outbreaks with the same *R*_0_ can have very different spatial spread (see Figure 3), *R*_0_ is still a useful variable to predict whether an epidemic will die out or keep growing.

### Outbreaks across regional boundaries

In order to complement the mapping out of the phase diagram, the random-walk outbreak model was also used to simulate real-world infectious dynamics. In this section, the effects of having multiple regions with separate *p* values (representing adjacent states or provinces with different distancing or shelter-in-place policies) is explored. The spatial aspect of this scenario is simplified to two infinite half-planes with distinct *p* values.

Most notably, the random-walk model corroborates the hypothesis that it is possible for an infection to grow very slowly (constantly on the brink of completely dying out) until it eventually reaches a region with a higher value of *p*, upon which it exhibits rapid growth. This is demonstrated in Figure 5. Moreover, there is also a significant backflow of infectors returning from the high-*p* region to the original low-*p* region.

**Figure 5.**
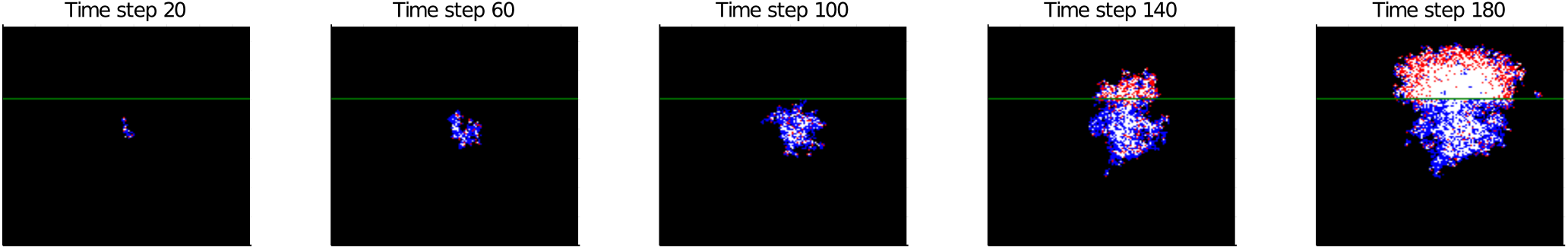
Simulation of an outbreak spanning two regions with distinct *p* values. The border between the two regions is located at the green *y* = 100 line and the infection originates at the center of the grid. Once crossing the boundary to the upper region, the infection explodes. For *y ≤* 100, *p* = 0.1 and for *y* > 100, *p* = 0.3. This illustrates a stark example of the effects of an infection re-igniting upon crossing a regional border. The boxes are 150 × 150 sites.

The initial states of epidemics near a regional boundary can be described by the following model. Consider two regions separated by a linear boundary with the infection probabilities *p*_1_ and *p*_2_ correspondingly. Suppose a random walker starts near the border, and its walk is not affected by the border itself (e.g., no checkpoints on boundary). Then the ratio of the numbers of infected persons in regions 2 and 1 is (see Methods):

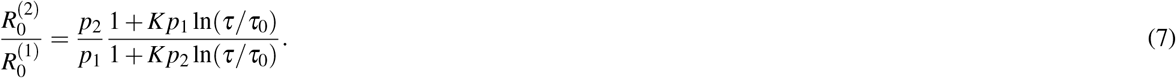

where *K* and *τ*_0_ are the same as in equation (4). For short walks (*K* max(*p*_1_, *p*_2_) ln(*τ/τ*_0_) ≪ 1) this expression becomes *p*_2_*/p*_1_, which corresponds to the “naïve” idea that the outbreak sizes in each region are proportional to *p*. However for long walks when *K* min(*p*_1_, *p*_2_) ln(*τ/τ*_0_) dominates, the ratio becomes 1, which means when the infection period is long enough, the infected agent revisits their contacts many times, hence the number of secondary infections no longer depends on the probability to infect during one meeting.

### Spatial characteristics and scaling of the outbreak

In our spatial model, we can also study the growth and geometry of the affected area and, in particular, its scaling in the critical region. To this end, we can consider in more detail one cross-section of the phase boundary of Figure 4. Fixing *τ* = 50, according to Eq. (4) the critical value of *p* is *p*_c_ ≈ 0.0283. We are interested in the behavior in the region of indefinite growth as we approach this limit, so we have carried out simulations for *p* = 0.03, 0.035, 0.04, 0.045, 0.05, 0.055, 0.06 (if we go below *p* = 0.03 too many runs die out, the statistics grows noisier and it takes too long to reach the asymptotic regime). For each *p* we followed 500 independent runs up to *t* = 1500 steps. The SM includes a collection of snapshots from these simulations for several values of *p*.

The first quantity that we can study is the growth of the number *R* of removed/recovered sites as a function of time. As expected, all of our simulations are in the indefinite-growth regime (see SM for lower values of *p*, where *R* plateaus as the epidemic dies out). The behavior with time seems to approach a power law, but is difficult to analyze. We can, however, make use of the spatial nature of our model to find a more appropriate scaling variable. Indeed, a simple and intuitive spatial observable is the radius of gyration ℛ_g_ of the cluster of removed sites. If **r**_*i*_ is the position of the *i*-th removed site, then

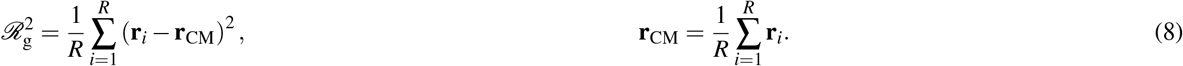

As we can see in Figure 6, in terms of this variable the growth of *R* rapidly approaches a power law. Moreover, the curves for all our values of *p* collapse. Performing a joint fit of all the points to 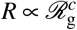 for ℛ_g_ ≥ 20, we obtain *c* = 2.04(2) with a goodness-of-fit metric of *χ*^2^*/*d.o.f. = 12.27*/*30, where d.o.f. = number of degrees of freedom. In other words, despite the low value of *p*, the number of infected sites scales as a volume (i.e., a large proportion of sites get infected, see SM for a direct measure of this attack rate), so the clusters are not space-filling fractals. The infection clusters could be further characterized by analyzing the scaling of the number *S* of surface sites. A site is on the surface if it a) belongs to the connected cluster of removed sites growing from the origin and b) at least one of its nearest neighbors is susceptible. Surface estimations are, however, noisy and possibly affected by stronger scaling corrections. Indeed, in this case only the curves for 0.03 ≤ *p* ≤ 0.04 (those closest to the critical point) collapse, yielding an exponent *c*_*S*_ = 1.92(5), with *χ*^2^*/*d.o.f. = 8.25*/*8 (see SM for figures). This exponent is not significantly different from *c* _*S*_ = 2, which would correspond to a linearly widening surface layer, but in any case its value is not precise enough to determine the universality class of the problem. Indeed, as we show in the next section, *S* is not the most appropriate observable for such a determination.

**Figure 6.**
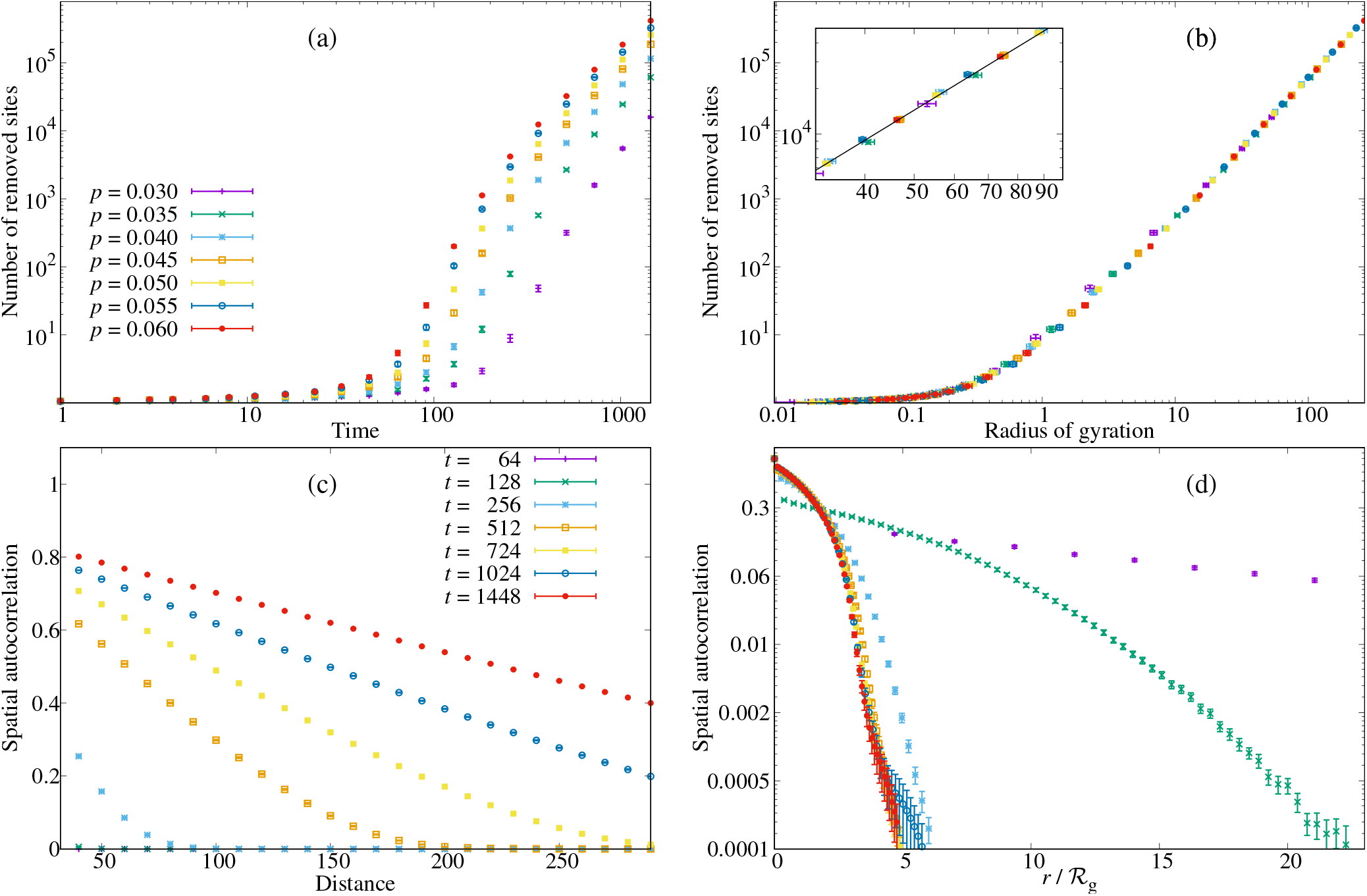
Spatial growth of the outbreak with time. (a) Total number of removed sites (total epidemic size) as a function of time in a log-log scale. (b) Same quantity as a function of the radius of gyration. The inset shows a closeup to appreciate the power-law fit. (c) Normalized spatial autocorrelation of the set of removed sites, *ρ* in Eq. (9), for *p* = 0.05. (d) Spatial autocorrelation in semi-log scale as a function of *r/*ℛ_g_. Again, using ℛ_g_ as scaling variable leads to an enveloping curve.

Finally, we can characterize the shape of the outbreak by studying its spatial autocorrelation function. For all the sites in the lattice, we define a function *g*(**x**) that is 1 if the site is removed and 0 otherwise. Then

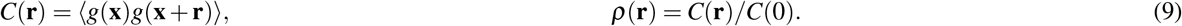

We have plotted *ρ* for *p* = 0.05 in panels (c) and (d) of Figure 6 (see other *p* in the SM). Measuring the distance in units of ℛ_g_ again leads to an enveloping curve as time grows, confirming that this is an appropriate length scale for the system.

Notice that our approximately quadratic growth of *R* with ℛ_g_ translates into a power-law, rather than exponential, incidence of the epidemic in time, as observed in recent works^17–19^ (ℛ_g_ itself approaches a power law in time with an exponent close to 1, see SM).

### Universality class of the problem and directed percolation

A natural conjecture regarding a random, *d*-dimensional branching process with an absorbing state (recovered/removed) is that it falls into the (*d* + 1)-dimensional directed-percolation universality class. If this is the case, scaling at the critical point between finite and indefinite growth is described by three independent exponents: *β, ν*_⊥_ and *ν*_‖_. For *d* = 2, we have *β* = 0.583(3), *ν*_⊥_ = 0.733(8) and *ν*_‖_ = 1.295(6).^20, 21^

In the scaling regime, the size (mass or number of sites) of the growing space-time cluster on transverse (two-dimensional) length scale *ℓ*_⊥_ and longitudinal (time) scale *ℓ*_‖_ is given by

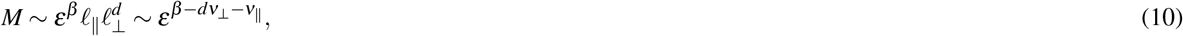

where *ε* is the distance from criticality in the control parameter. Eq. (10) assumes that lengths scale like the corresponding correlation lengths along the different axes, e.g., 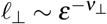.^22, 23^

Expressing the total space-time size of the epidemic in terms of the transverse-space scale only yields

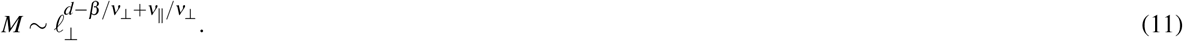

This is not, however, the scaling within a space-like slice (i.e., the clusters we analyzed in the previous section). One way to find that spatial scaling is to treat the exponent in Eq. (11) as a fractal dimension and intersect the epidemic’s space-time history with a transverse slice. The intersection set will scale with the transverse scale *ℓ*_⊥_ with exponent

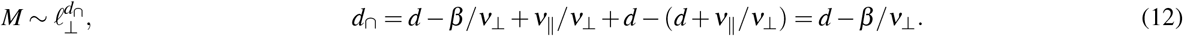

For (2 + 1)-dimensional directed percolation we obtain *d*_∩_ = 1.20(1). In other words, the number of active sites at fixed time should scale like *N*_active_ ∼ *ℓ*^1.20^ as a function of a typical spatial scale *ℓ*. Translating this analysis back into the language of epidemiology, the analogy with directed percolation predicts that the number of infected individuals *I* in a cluster of radius of gyration ℛ_g,*I*_ scales as 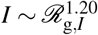 — a law that is measurably different from the Euclidean picture of a compact cluster with an active boundary of radius ℛ_g,*I*_.

We have tested this theory in Figure 7, by plotting the total number of currently infected sites as a function of the radius of gyration of this set. In this case, unlike for the number *R* of removed sites, the curves for different *p* do not collapse. In the critical region of 0.03 ≤ *p* ≤ 0.04, however, we can compute fits to 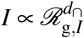. Averaging the resulting values of the exponent, we obtain *d*_∩_ = 1.18(5), compatible with the above prediction and clearly different from the mean-field prediction of *d*_∩_ = 1.

**Figure 7.**
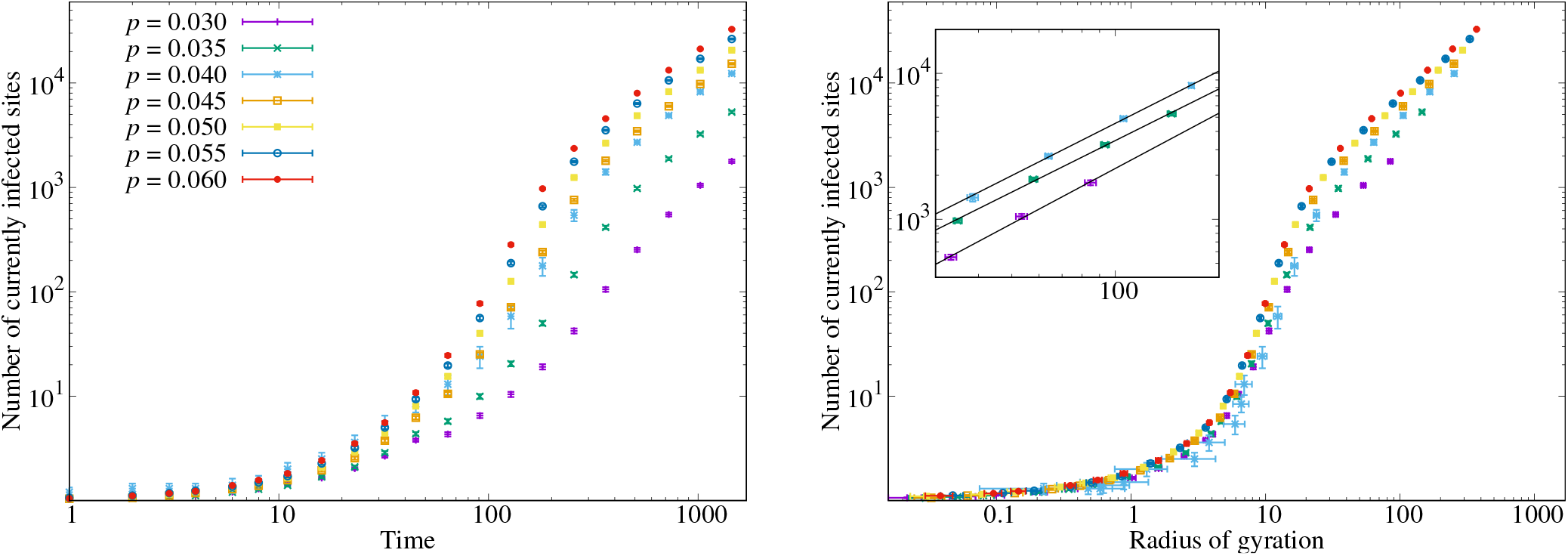
Growth of the number of currently infected sites, against time (left) and against the radius of gyration of the set (right). In this case the curves for different *p* do not collapse, but the curves approach a power law 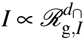 with *d*_∩_ = 1.18(5), see Eq. (12). This exponent is compatible with the directed-percolation universality class. The inset shows a closeup of the fits for *p* = 0.03, 0.035, 0.04.

## Discussion

We see that a simple lattice model provides a number of interesting insights about epidemic spread. One of the most useful features of idealized models like this one is that they provide an understanding of which factors are salient for the outcome, and which are not. This understanding can be used in the construction of more detailed and realistic models, in the same way that a pencil sketch is useful when starting a full-size oil painting.

One of the most important questions when studying epidemics is whether the value of *R*_0_ is sufficient to describe the epidemics behavior, or whether a detailed analysis of the contact network is necessary. Our model shows that the answer depends on what aspect of the epidemic is interrogated. The spatial features of the outbreak are not determined by *R*_0_ alone, but depend on the individual infectivity and the number of individual contacts separately. In contrast, the phase boundary between infinite spread and localization of the epidemic is still determined by *R*_0_.

We provide the dependence of *R*_0_ on the easily interpretable and intuitively clear parameters: the average number of steps per infected person, the probability to infect another person, the average length of a step. This information can be used to form policy decisions. For example, we find that the step length is important if the probability to infect is high enough (above we give the estimate *Kp*ln(*τ/τ*_0_) ≫ 1 for the threshold value of *p*). This is relevant to the travel restriction, especially as more contagious strains are discovered.

Finally, we have studied the scaling of our outbreaks in terms of space and length scales in the critical region between the extinction and the indefinite spreading regime. By analyzing the fractal nature of the growing cluster, we make a quantitative connection to the problem of directed percolation, whose universality class seems to rule our critical regime.

A significant idealization in our model is the motion of the infective agents: the random walkers can go anywhere on the lattice and do not have a memory of their origins. In real life, people tend to have permanent residences and return there daily. These periodic movements would work to make the outbreaks even more localized than those in our model. Introducing more realistic walking patterns is, therefore, a compelling direction for future work. Another interesting possibility would be to use the end state of one simulation, with a lattice including both removed and still susceptible sites, as the initial condition to simulate the effect of subsequent epidemic waves. Such a numerical experiment has a direct bearing on the question of thresholds to herd immunity in a fully spatial model, a topic that has received scant attention.

## Methods

### Effect of fluctuations on phase diagram

The phase boundary introduced above separates the cases where an outbreak dies out from those where it spreads indefinitely. Let *P*_0_ be the probability that the outbreak dies out given one index case. We assume that the number of people infected by the index case has a Poisson distribution with mean *R*_0_. The meaning of *R*_0_ is thus the same as in classical epidemiology: the number of people infected by the index case in a naïve population.^2^ The probability that the number of secondary infections is zero, and that the outbreak dies out immediately, is exp(−*R*_0_). The probability that the index case infects exactly *n* persons is 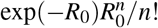 If the index case infected *n* persons, the epidemic dies out only if each secondary outbreak caused by each of these *n* persons dies out. The probability of such an event *P*_1_ is, strictly speaking, different from *P*_0_ since the secondary infections operate in a different environment than the index case. However, we neglect this and stipulate that *P*_1_ ≈ *P*_0_. The probability that *n* secondary outbreaks die out is therefore 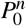. Then we can write down

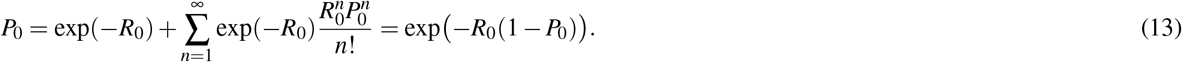

At 0 ≤ *R*_0_ ≤ 1 the only solution for this equation is *P*_0_ = 1. This means that at *R*_0_ ≤ 1 the epidemics is always localized. At *R*_0_ > 1 equation (13) gets a solution between 0 and 1. If we draw the phase boundary at the point where exactly 50% of outbreaks survive, then the critical value of *R*_0_ is *R*_0_ = 1.39. This agrees extremely well with Figure 4.

If we have *N* index cases, the probability that the epidemic is localized is, to first approximation, 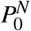. At large *N* this probability quickly approaches zero, for *R*_0_ > 1. It is important to note that the phase boundary, unlike the other features of the epidemic, depends only on *R*_0_. Note that this derivation does not use the lattice approximation and thus should be valid for a wide class of models. The only crucial assumption for the derivation is that the environment for the secondary infections is the same as for the index case. This assumption is true for well-mixed models and becomes less good in the case of high spatial inhomogeneity around the index case.

### Asymptotics for *R*_0_

In this section, we derive a theoretical estimate for *R*_0_: the number of infections produced by one random walker in a susceptible population. We use ideas similar to those in^24–26^.

Let us consider a walker that starts at the point **x**. Let *C*(*τ, m*, **x, w, y**) be the probability for this walker to end up at the point **y** after making *τ* steps, while visiting the point **w** exactly *m* times. The probability that the walker does *not* infect the site **w** during this trip is (1 − *p*)^*m*^, since the probability to infect at each visit is *p*. Summing over **w, y** and *m*, we get the total number of infected sites as

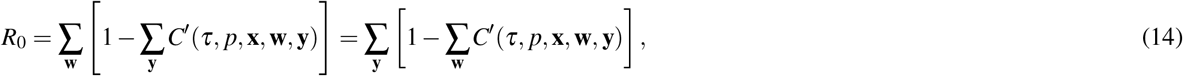

where

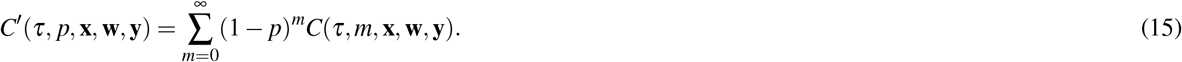

This function is (up to a normalization) the generating function introduced in Rudnick and Gaspari [26, Section 3.2]. To calculate it, we introduce a transformation similar to the transition from the canonical ensemble to the grand canonical ensemble in statistical physics. Namely, let us take a complex variable *z* and write down the series:

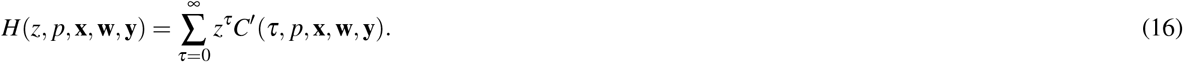

The inverse transformation is

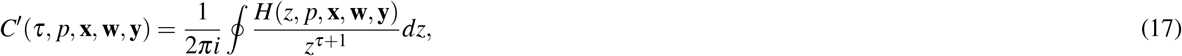

where the complex integral is taken around the point *z* = 0. As we shall see below, the value of this integral is defined by the singularity of *H* at *z* → 1.

Let *P*(*τ*, **x, y**) be the probability for a random walker starting at the point **x** to end at the point **y**. We introduce two functions

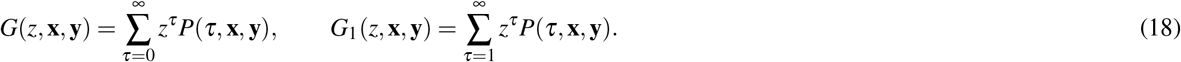

A remarkable theorem [26, Section 3.3] states that function *H* can be expressed through the functions *G* and *G*_1_:

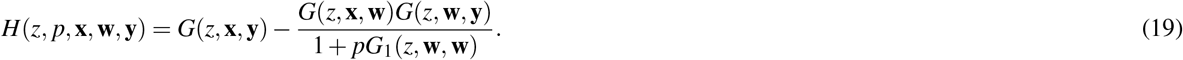

Integration of *G* over *y* and *w* is easy due to the normalization. The only nontrivial contribution is from the function *G*_1_ in the denominator of this expression. In two dimensions, this function can be expressed as [26, Section 3.5]

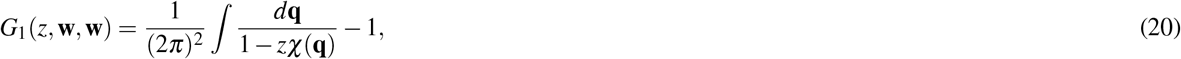

where *χ*(**q**) is the structure factor, *i*.*e*. the Fourier transform of the jump probability function. More precisely, let *p*(**x**) be the probability for the walker to go from the point **0** to the point **y** in one step. We then define the structure factor *χ*(**q**) as

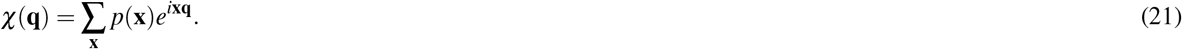

At small *q*, we can expand the structure factor as

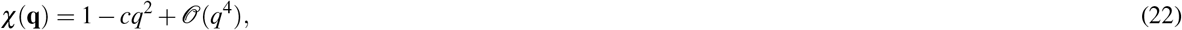

which gives near the singularity *z* → 1

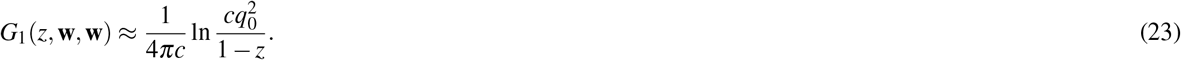

Here *q*_0_ ≈ 1 is the upper limit in the integral (20) (assuming the lattice constant to be 1). Using these asymptotics, and integrating equations (14), (17), and (19), we get equation (4) with

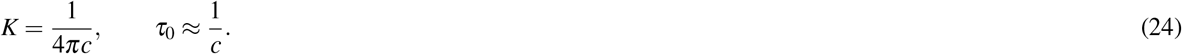

Note that we use a different normalization from the one used in the Rudnick and Gaspari^26^: they count the number of random walks, whereas we count the probability. This means that the singularities of the functions *H, G* and *G*_1_ are at *z* = 1 instead of *z* = *z*_*c*_, and that *χ*(0) = 1.

The coefficients *K* and *τ*_0_ depend only on the structure factor *χ*. To calculate these two, let us generate a large number *N* of jumps according to equations (2) and (3). Let us take **q** along the *x* axis. If jump number *i* lands at the point **r**_*i*_ = (*x*_*i*_, *y*_*i*_), then it contributes the term

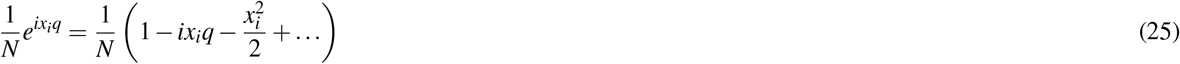

to *χ*. Due to symmetry, the contribution of the second term is zero, and we arrive at

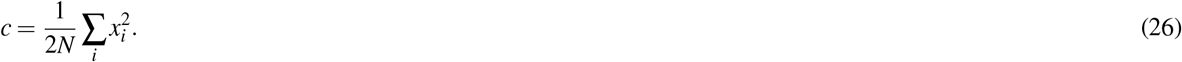

In our experiments, we generate *N* = 10^6^ points, which gives *c ≈* 0.458, and the resulting values of *K* and *τ*_0_ in equation (5).

### *R*_0_ in inhomogeneous case

The method described in the previous section can be easily generalized for the case when the probability of infection *p* is different for different sites. Indeed, in equation (15) *C*′ depends only on the value *p*_**w**_ in the point **w**. Therefore we can take the expressions for *C* from that section, calculate *C*′, and obtain *R*_0_ by integration in equation (14). Let us consider two semi-infinite regions separated by a linear border with the probabilities of infection *p*_1_ and *p*_2_. Suppose a walker starts in the first region at the distance *d* from the border. Then integration in equation (14) gives

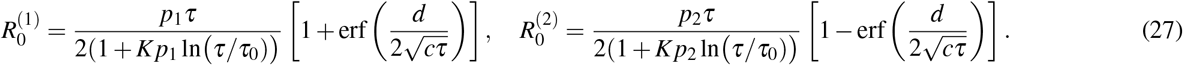

At *d* ≪ *c*^2^*τ*^2^ we get equation (7).

## Data Availability

Code and data for figures and tables can be found in the linked GitHub repository.

https://github.com/czbiohub/random-walk-epidemic-model

## Data availability

Code and data for figures and tables can be found at https://github.com/czbiohub/random-walk-epidemic-model.

## Acknowledgments

We are grateful to the anonymous reviewers for their probing questions and deep insights. A.C., G.H., A.M., and D.Y. are supported by the Chan Zuckerberg Biohub; B.V. is supported by the Chan Zuckerberg Initiative. D.Y. acknowledges support by MINECO (Spain) through Grant No. PGC2018-094684-B-C21, partially funded by the European Regional Development Fund (FEDER).

## Author contributions statement

G.H. and A.M. designed research; A.C. performed simulations with guidance from G.H., A.M. and D.Y.; A.M. wrote the simulation code; B.V. carried out random-walk analysis; all authors analyzed data and contributed to the writing of the paper.

